# Evaluating equitable care in the ICU: Creating a causal inference framework to assess the impact of life-sustaining interventions across racial and ethnic groups

**DOI:** 10.1101/2023.10.12.23296933

**Authors:** Tristan Struja, João Matos, Barbara Lam, Yiren Cao, Xiaoli Liu, Yugang Jia, Christopher M. Sauer, Helen D’Couto, Irene Dankwa-Mullan, Leo Anthony Celi, Andre Kurepa Waschka

## Abstract

**Background:** Variability in the provision of intensive care unit (ICU)-interventions may lead to disparities between socially defined racial-ethnic groups.

**Research Question:** We used causal inference to examine the use of invasive mechanical ventilation (IMV), renal replacement therapy (RRT), and vasopressor agents (VP) to identify disparities in outcomes across race-ethnicity in patients with sepsis.

**Study Design and Methods:** Single-center, academic referral hospital in Boston, Massachusetts, USA. Retrospective analysis of treatment effect with a targeted trial design categorized by treatment assignment within the first 24 hours in the MIMIC-IV dataset (2008- 2019) using targeted maximum likelihood estimation. Of 76,943 ICU stays in MIMIC-IV, 32,971 adult stays fulfilling sepsis-3 criteria were included. The primary outcome was in-hospital mortality. Secondary outcomes were hospital-free days, and occurrence of nosocomial infection stratified by predicted mortality probability ranges and self-reported race-ethnicity. Average treatment effects by treatment type and race-ethnicity, Racial-ethnic group (REG) or White group (WG), were estimated.

**Results:** Of 19,419 admissions that met inclusion criteria, median age was 68 years, 57.4% were women, 82% were White, and mortality was 18.2%. There was no difference in mortality benefit associated with the administration of IMV, RRT, or VP between the REG and the WG. There was also no difference in hospital-free days or nosocomial infections. These findings are unchanged with different eligibility periods.

**Interpretation:** There were no differences in the treatment outcomes from three life-sustaining interventions in the ICU according to race-ethnicity. While there was no discernable harm from the treatments across mortality risk, there was also no measurable benefit. These findings highlight the need for research to understand better the risk-benefit of life-sustaining interventions in the ICU.

## INTRODUCTION

Researchers have repeatedly discovered racial and ethnic disparities in critical illness and end-of-life care ^1,2^. An analysis of over 1,000 patients with a history of stroke showed that black patients in the cohort were less likely than white patients to use hospice and more likely to have multiple emergency department visits, hospitalizations, and intensive treatments in their last six months of life ^3^. While the hypothesized reasons for these types of findings are multifactorial, disparities in who receives life-sustaining treatments have also raised the question of subconscious and systemic biases ^1^. In a study of over 17,000 intensive care unit (ICU) admissions, white patients received more technological monitoring, laboratory testing, and life-supporting treatments compared to black patients on the first day of their ICU stay ^4^. Another study of over 28 hospitals in the United States showed that black patients with pneumonia were less likely to receive guideline-adherent antibiotics and more likely to receive mechanical ventilation ^5^. These disparities were highlighted during the COVID-19 pandemic when researchers found that there were not only differences in interventions across racial and ethnic groups ^6^, but also in survival outcomes ^7,8^.

Uncovering disparities in care is more urgent than ever given the rising popularity of artificial intelligence (AI) technology. Now, disparities have the opportunity to cause harm twice: their existence can drive inequities in care today and perpetuate bias in AI algorithms tomorrow ^9^. Conducting randomized controlled trials (RCTs) to understand how life-sustaining interventions might lead to different outcomes across racial and ethnic groups is both unethical and impracticable. These types of research questions are best answered using observational data, but there are limitations to databases such as claims registries, which often lack important clinical details ^10,11^. With the development of high-resolution datasets such as MIMIC-IV, we can apply a causal inference framework to leverage this real-world data in understanding how different patients have been affected by different interventions ^12^.

In this study, we provide a causal inference framework for assessing the impact of life-sustaining interventions across different racial and ethnic groups in the MIMIC-IV database. MIMIC-IV is one of the most widely used datasets in both critical care and machine learning research; it is therefore imperative that we understand what potential biases exist in the data. We also provide the code for our framework so that other groups can readily assess for potential inequities in critical care at their own organizations.

## METHODS

This study is reported in accordance with the Strengthening the Reporting of Observational studies in Epidemiology (STROBE) statement ^13^. The language of this paper follows the American Medical Association’s reccomendations ^14^. Data were extracted from the open-access and de-identified MIMIC-IV using Google’s BigQuery software. MIMIC is maintained by the Laboratory for Computational Physiology at Massachusetts Institute of Technology (MIT) ^15^. Approval for the study and a waiver of consent was obtained from the Institutional Review Boards as all the data is de-identified. MIMIC-IV includes physiologic data collected from bedside monitors, as well as other clinical variables and provider documents recorded in the ICU electronic medical record. Approximately 70,000 de-identified medical ICU records are archived in MIMIC-IV.

We hypothesized that treatment allocation of ICU interventions is not equally distributed across race-ethnicity leading to differences in outcomes. We suspected that Racial-ethnic patient group with sepsis experienced a more harmful use of ICU interventions, especially in less severely ill patients.

### Cohort Selection

All patients older than 18 years of age who had sepsis as defined by the sepsis-3 criteria were included in the analysis ^16^. We only included first-time ICU stays, and excluded cases missing race or ethnicity information or discharge location. This also includes patients with race “other” as this category can comprise patients of all ethnicities. Patients were excluded if they had a length of stay (LOS) of less than one day or more than 30 days to ensure a homogenous cohort. To avoid immortal time bias, treatment assignment was only possible during an eligibility period of the first 24 hours for invasive mechanical ventilation (IMV) and vasopressors (VP), and first three days for renal replacement therapy (RRT). Immortal time bias occurs in observational studies when there is failure to align start of follow-up, specification of eligibility, and treatment assignment leading to a time window during the study where, for inclusion in the cohort, the patients are required to be *immortal* to the event of interest ^17^. It can lead to erroneous conclusions about the effectiveness of treatments or interventions if not properly accounted for in the study design and analysis. If treatment was started after this eligibility period the patient was retained in the control group, emulating a targeted trial (see **eTable 1**).

### Covariates

Patient-level variables were obtained from the database at the time of ICU admission and time-varying variables were aggregated for the first 24 hours of the stay by taking the maximum, minimum, or mean value as appropriate (see **eTable 2**). We further extracted ICD-10 codes for key comorbidities, including hypertension, COPD, asthma, heart failure, stages of chronic kidney disease (CKD), and Oxford Acute Severity of Illness Score (OASIS). Patients with a label for White (e.g., White, White – Brazilian, White – Russian) were grouped as White (WG), while we grouped the remainder as Racial-ethnic patient group (REG) as race-ethnicity is heavily imbalanced in MIMIC (i.e., approximately 80% White patients).

ICU admission OASIS scores were used to calculated predicted mortality probability (PMP) to give an easily generalizable and externally valid scale ^18–20^. Patients were categorized in PMP quartiles as having low, moderate, high, or very high sepsis severity.

### Outcomes

The primary outcome was in-hospital mortality, including discharge to hospice care. Secondary outcomes were hospital-free days by day 28 and combined nosocomial infection (central-line associated bloodstream infection, catheter associated urinary tract infection, surgical site infection, and ventilator associated pneumonia). In line with recent research, we used the time of discharge or death at an odd versus an even hour as a negative control outcome ^21^, as treatment should not affect this random event.

### Statistical Analysis

Statistical analysis was performed using *R* version 4.2 ^22^ and *Python* 3.10 ^23^ running on Visual Studio Code. We used targeted maximum likelihood estimation (TMLE) models adjusted for confounders to compute the average treatment effect (ATE) for each of the interventions, stratified by race-ethnicity and predicted mortality range ^24^. TMLE is a semiparametric framework estimating the causal effect of an intervention on an outcome of interest. For counterfactual modelling, we used the *SuperLearner* package, an ensemble machine learning algorithm with 5- fold cross-validation. The ATE is a marginal effect, meaning it is averaged over the covariates. ATE is defined as the difference in outcome should everyone be treated with the invasive treatment versus no one being treated, as shown in equation 1:

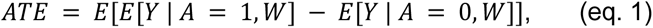

Where Y represents the outcome (primary: in-hospital mortality; secondary: hospital-free days by day 28 and combined nosocomial infections; negative control: death or discharge at odd hour); A the treatment (IMV, RRT, or VP); and W the confounders for adjustment.

Treatment assignment was assumed to be independent of the outcome (i.e., conditional exchangeability assumption). There were no multiple versions of a treatment, as we dichotomized all treatments (i.e., consistency assumption). We tabulated the treatments according to race-ethnicity and predicted mortality range to check whether all strata were eligible to receive a treatment (i.e., positivity assumption).

In a sensitivity analysis, we repeated the calculations changing the eligibility period for RRT from 3 days to 1 day or 5 days. We consider our work hypothesis generating, which is why we abstained from computing p-values and only provided 95% confidence intervals.

## RESULTS

After applying our inclusion and exclusion criteria, we were left with a sample size of 19,419 admissions (see **Figure 1**). **Table 1** shows the baseline characteristics of our cohort stratified by race-ethnicity. Median age was 68 years, 57.4% were women, 82% were White. Median length of stay (LOS) was 9.5 days and did not differ substantially if patients survived or died. 18.2% of patients admitted to an ICU died or were discharged to a hospice (2.7% of all discharges). Baseline Charlson comorbidity index (CCI), IMV, and VP use, OASIS and SOFA scores between races and ethnicities were evenly balanced. However, there were marked differences in the use of RRT (11.2% in the REG, 6.1% in the WG), in prevalence of hypertension, CKD, and age (median age 64.5 in the REG, 69 in the WG). All race-ethnicity groups and treatments had a non-zero probability of occurring in each of the predicted mortality categories showing no indication for violation of the positivity assumption (see **eTable 3**).

**Figure 1:**
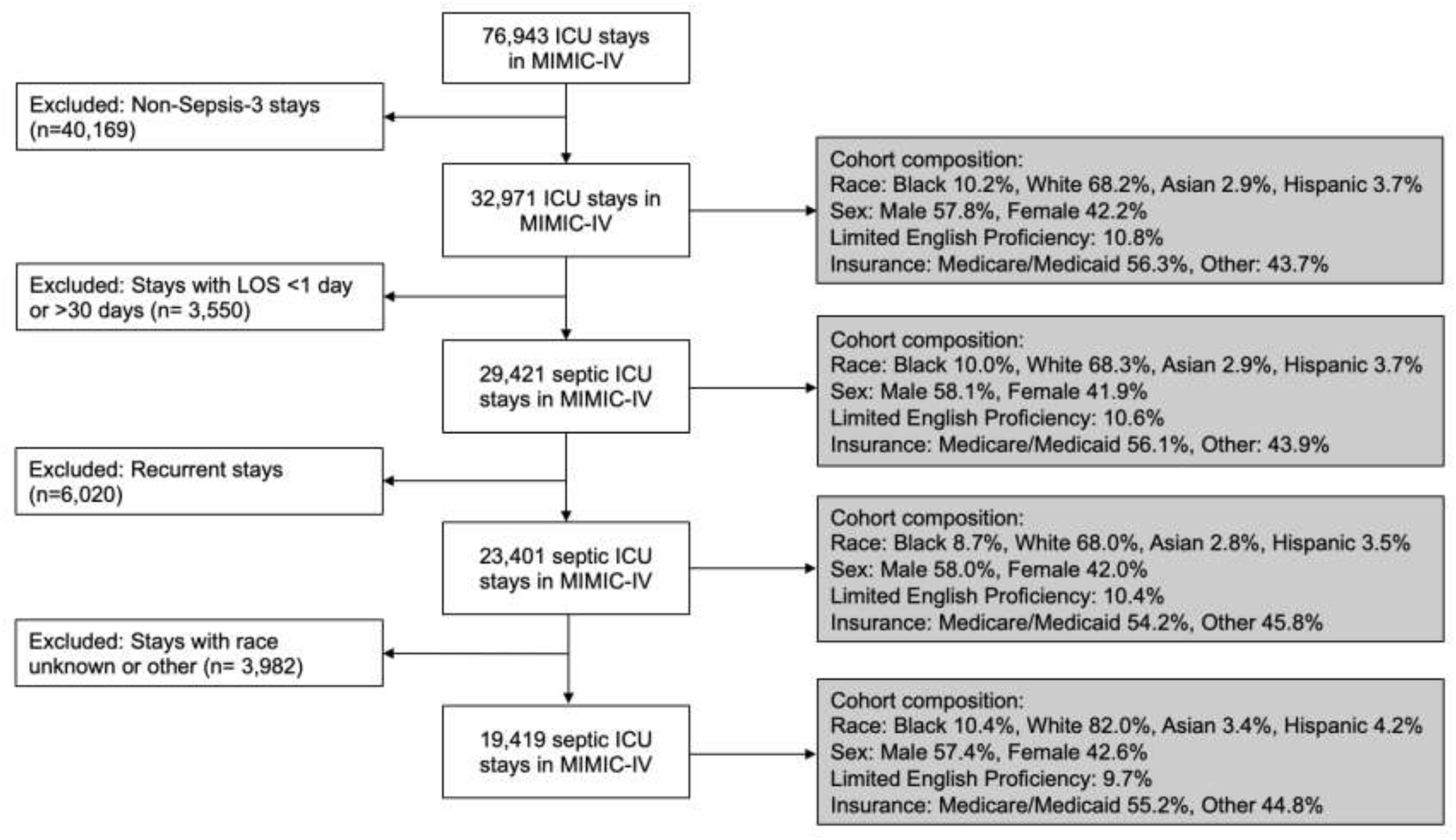
Study cohort selection flow chart. Abbreviations: LOS, Length of stay

**Table 1:** Baseline information on the study cohort.

|  | Racial-ethnic group<br>(N=3,496) | White group<br>(N=15,923) | Overall<br>(N=19,419) |
| --- | --- | --- | --- |
| **Racial-ethnic group** |  |  |  |
| Asian | 659 (18.8%) | 0 (0%) | 659 (3.4%) |
| Black | 2,027 (58.0%) | 0 (0%) | 2,027 (10.4%) |
| Hispanic | 810 (23.2%) | 0 (0%) | 810 (4.2%) |
| White | 0 (0%) | 15,923 (100%) | 15,923 (82.0%) |
| **In-hospital mortality** | 664 (19.0%) | 2,866 (18.0%) | 3,530 (18.2%) |
| **Discharge to hospice** | 102 (2.9%) | 417 (2.6%) | 519 (2.7%) |
| **Elective admission** | 325 (9.3%) | 2,740 (17.2%) | 3,065 (15.8%) |
| **Vasopressors started in eligibility period** | 1,337 (38.2%) | 7,299 (45.8%) | 8,636 (44.5%) |
| **Mechanical ventilation started in eligibility period** | 1,514 (43.3%) | 7,036 (44.2%) | 8,550 (44.0%) |
| **Renal replacement therapy started in eligibility period** | 390 (11.2%) | 974 (6.1%) | 1,364 (7.0%) |
| **Central line-associated bloodstream infection** | 3 (0.1%) | 11 (0.1%) | 14 (0.1%) |
| **Catheter-associated urinary tract infection** | 4 (0.1%) | 17 (0.1%) | 21 (0.1%) |
| **Surgical site infection** | 11 (0.3%) | 45 (0.3%) | 56 (0.3%) |
| **Ventilator-associated pneumonia** | 23 (0.7%) | 60 (0.4%) | 83 (0.4%) |
| **Nosocomial infections combined** | 39 (1.1%) | 125 (0.8%) | 164 (0.8%) |
| **Age categorical (years)** |  |  |  |
| 18 - 44 | 514 (14.7%) | 1,206 (7.6%) | 1,720 (8.9%) |
| 45 - 64 | 1,234 (35.3%) | 4,943 (31.0%) | 6,177 (31.8%) |
| 65 - 74 | 756 (21.6%) | 3,950 (24.8%) | 4,706 (24.2%) |
| 75 - 84 | 638 (18.2%) | 3,600 (22.6%) | 4,238 (21.8%) |
| 85 and higher | 354 (10.1%) | 2,224 (13.9%) | 2,577 (13.3%) |
| **Age overall (years)** |  |  |  |
| Median (IQR) | 64.5 (52.0, 76.0) | 69.0 (59.0, 79.0) | 68.0 (57.0, 79.0) |
| **Sex** |  |  |  |
| Female | 1,808 (51.7%) | 9,331 (58.6%) | 11,139 (57.4%) |
| Male | 1,688 (48.3%) | 6,592 (41.4%) | 8,280 (42.6%) |
| **OASIS** |  |  |  |
| Median (IQR) | 32.0 (27.0, 39.0) | 32.0 (27.0, 38.0) | 32.0 (27.0, 38.0) |
| **SOFA score** |  |  |  |
| Median (IQR) | 5.00 (3.00, 8.00) | 5.00 (3.00, 8.00) | 5.00 (3.00, 8.00) |
| **Length of stay (days)** |  |  |  |
| Median (IQR) | 9.73 (5.77, 16.8) | 9.06 (5.71, 15.6) | 9.54 (5.72, 15.7) |
| **Length of stay, if survived (days)** |  |  |  |
| Median (IQR) | 9.77 (5.85, 17.0) | 8.83 (5.76, 15.5) | 9.56 (5.78, 15.6) |
| **Length of stay, if died (days)** |  |  |  |
| Median (IQR) | 9.27 (4.74, 16.2) | 9.41 (4.72, 16.6) | 9.38 (4.72, 16.6) |
| **Charlson comorbidity index categorical** |  |  |  |
| 0 - 3 | 752 (21.5%) | 2,966 (18.6%) | 3,718 (19.1%) |
| 4 - 6 | 1,161 (33.2%) | 6,587 (41.4%) | 7,748 (39.9%) |
| 7 - 10 | 1,279 (36.6%) | 5,338 (33.5%) | 6,617 (34.1%) |
| 11 and above | 304 (8.7%) | 1,032 (6.5%) | 1,336 (6.9%) |
| **Charlson comorbidity index continuous** |  |  |  |
| Median (IQR) | 6.00 (4.00, 8.00) | 6.00 (4.00, 8.00) | 6.00 (4.00, 8.00) |
| **Hypertension present** | 2,424 (69.3%) | 10,480 (65.8%) | 12,904 (66.5%) |
| **Congestive heart failure present** | 1,229 (35.2%) | 5,420 (34.0%) | 6,649 (34.2%) |
| **COPD present** | 809 (23.1%) | 3,804 (23.9%) | 4,613 (23.8%) |
| **Asthma present** | 47 (1.3%) | 214 (1.3%) | 261 (1.3%) |
| **Chronic kidney disease stage $\geq 3$**<br>**present** | 670 (19.2%) | 1,665 (10.5%) | 2,335 (12.0%) |

Legend: Racial-ethnic patient group includes Asian, Black, Hispanic, Latino, etc.
Abbreviations: IQR; interquartile range; COPD, chronic obstructive pulmonary disease; OASIS, Oxford Acute Severity of Illness Score; SOFA, sequential organ failure assessment

### Unadjusted Analysis: Primary Outcome

We started by analyzing the unadjusted probability of a patient receiving a treatment, should they died or survived, to identify initial patterns in the data and check consistency of TMLE analysis. In patients receiving IMV and VP, the proportion of patients to survive the treatment was consistently higher than dying on treatment, albeit differences diminished with increasing illness severity (see **Figure 2**). In patients receiving RRT, the proportion for patients surviving with treatment was consistently lower than dying on treatment. These trends were consistent when stratifying by race-ethnicity (see **eFigure 1**).

**Figure 2:**
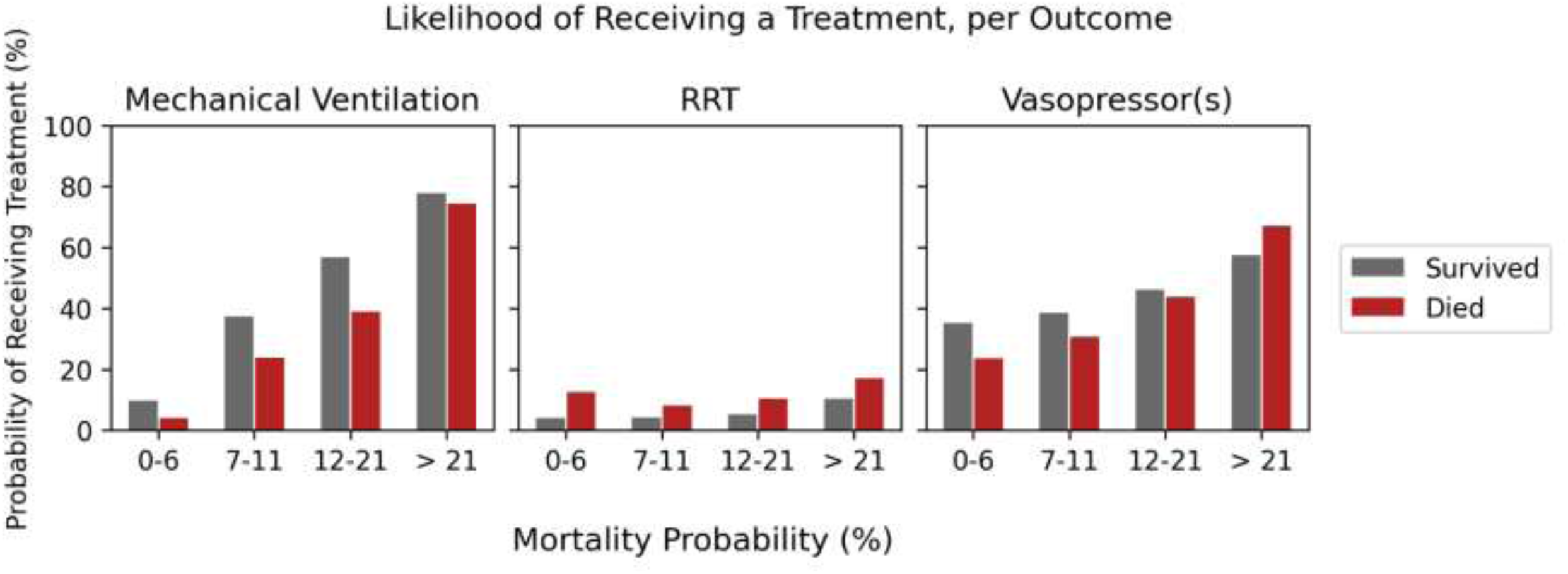
Distribution of patients across predicted mortality ranges, per invasive treatment. Abbreviations: RRT, renal replacement therapy

### TMLE: Negative Control Outcome

In adjusted TMLE modeling, there was no treatment effect in any intervention when discharge or death at an odd hour was used as a negative control outcome (see **eTable 4** and **Figure 3**). This suggests that residual confounding has been mitigated in our study ^21^.

**Figure 3:**
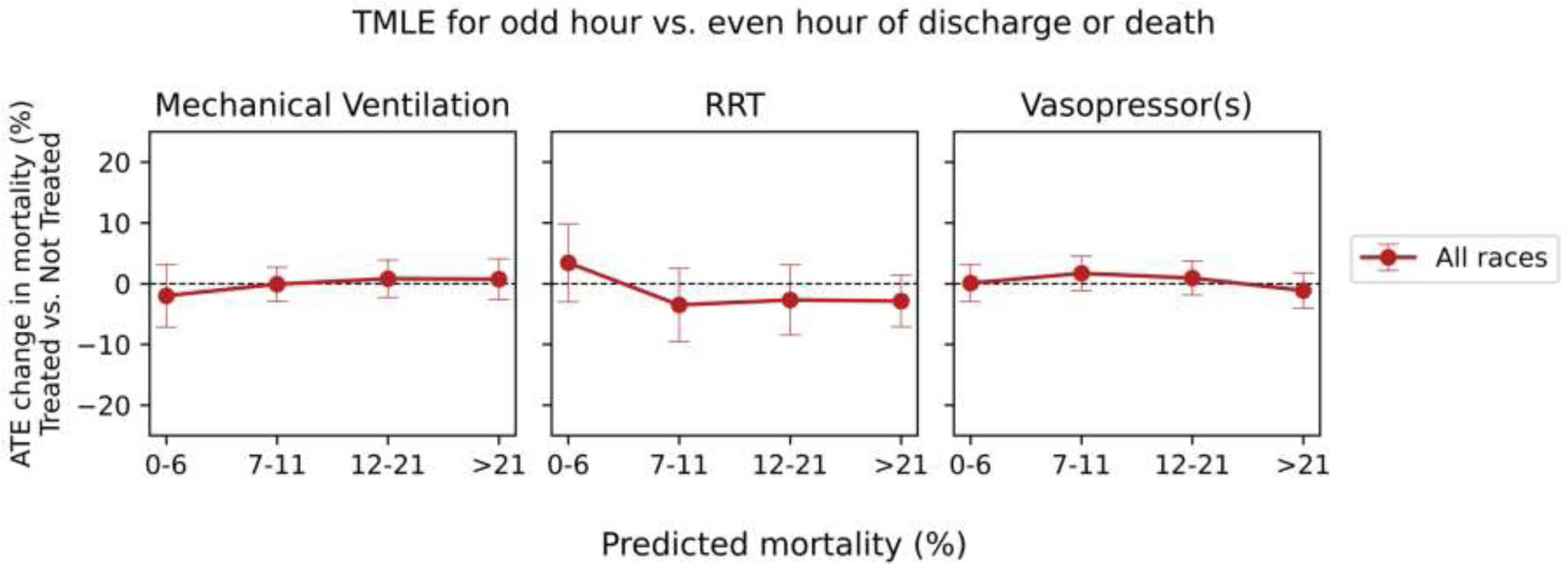
TMLE derived average treatment effects for change in probability of discharge or death at an odd hour vs. even hour of any given day as negative control outcome over predicted mortality categories. Abbreviations: ATE, average treatment effects; TMLE, targeted maximum likelihood; RRT, renal replacement therapy

### TMLE: Primary Outcome

We saw no clear distinction between the two racial groups for in-hospital mortality in any of the treatments (see **Figure 4** and **eTable 4**). In patients receiving IMV and VP, the confidence intervals for the ATE were crossing the null, except for the WG in the predicted mortality range of 0-6% and 12-21%. For patients under RRT, there was a consistent signal for harm in the lowest predicted mortality range of 0-6% with an ATE of 6.0% (95% CI 5.0%-7.0%) in the WG and ATE of 5.0% (95% CI 2.0%-7.0%) in the REG, but not in higher predicted mortality ranges. Results were similar in sensitivity analysis with RRT eligibility periods of 1 day or 5 days instead of 3 days (see **eFigure 2**).

**Figure 4:**
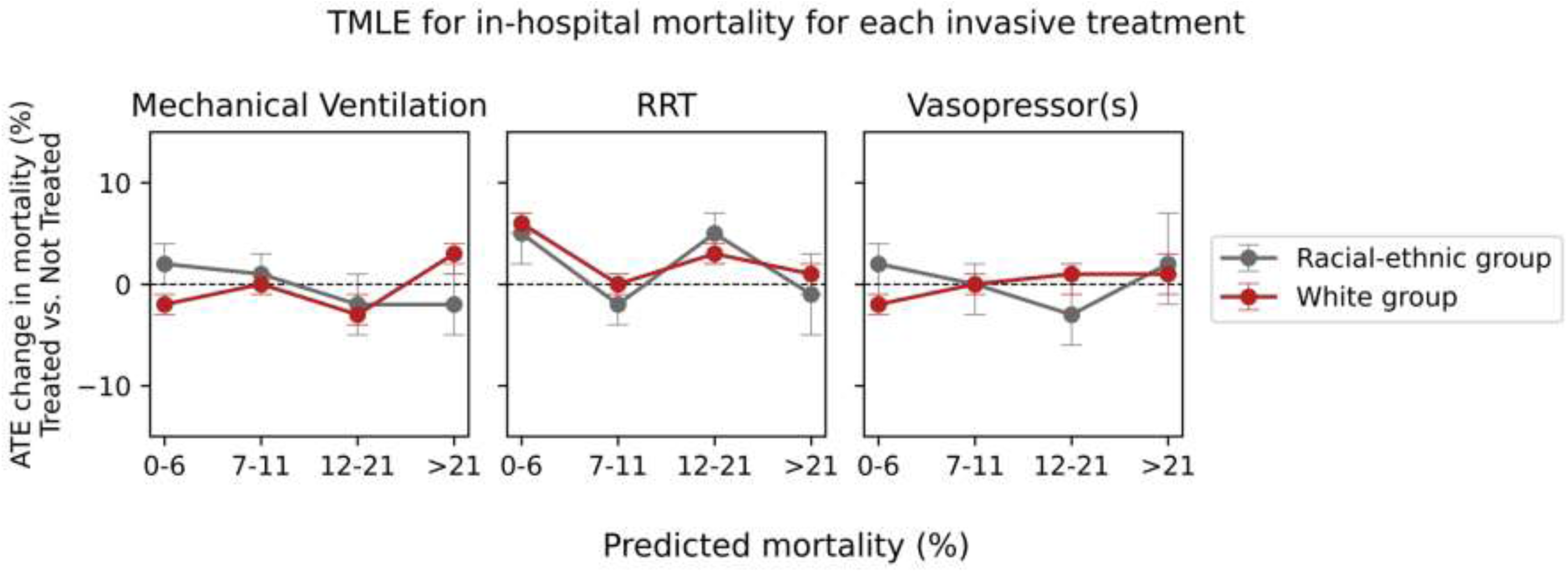
TMLE derived average treatment effects for change in probability of death over predicted mortality categories with RRT eligibility period of 3 days. Abbreviations: ATE, average treatment effects; TMLE, targeted maximum likelihood; RRT, renal replacement therapy

### TMLE: Secondary Outcomes

Hospital-free days were similar between the two racial groups, except for patients on RRT in the highest predicted mortality range >21% where we observed an ATE of 1.39 mean days (95% CI 0.66 to 2.12) in the REG and -0.4 mean days (95% -0.77 to -0.04) in the WG (see **eTable 4** and **eFigure 3**). Both groups had fewer hospital-free days when subjected to IMV, whereas VP did not influence the outcome. For the outcome of combined nosocomial infection, we consistently saw an ATE of 0% across all treatments and predicted mortality ranges.

## DISCUSSION

Using a causal inference framework to assess critically ill patients with sepsis in the MIMIC-IV database, we found no racial-ethnic disparities in the outcomes of patients treated with IMV, RRT, and VP. We also provide an open-source causal inference analytic pipeline that researchers can use to perform disparity research using electronic health record data.

Health equity is increasingly recognized by policymakers as essential to public health ^25,26^. In December of 2022, the Center for Medicare and Medicaid Services (CMS) proposed tying reimbursements to health equity outcomes ^27^, highlighting the importance of organizations having a way to *measure* health equity. The Centers for Disease Control and Prevention (CDC) cites measurement as one of the key pillars of advancing health equity, alongside population-level strategies and policy changes ^28^. Several frameworks have been proposed for describing the social determinants of health that lead to disparities ^29,30^, but organizations need an out-of-the-box tool they can easily and repeatedly use to assess measure health equity. Causal inference on high-resolution electronic health record data offers a robust method of detecting test and treatment disparities that are not explained by clinical factors and if present, quantifying their effect sizes on patient outcomes. We employed TMLE to investigate whether outcomes from IMV, RRT and VP vary across race-ethnicities after adjusting for time-varying clinical confounding, and our proposed framework is both free and easy to use, and can be run on a personal computer. TMLE is a semiparametric estimator offering several compelling properties. It is based on the targeted minimum loss-based estimation and machine learning algorithms to minimize the risk of model misspecification ^31^. As TMLE is doubly robust, if either the outcome regression or the exposure mechanism is consistently estimated, it will yield unbiased estimates, and when both the treatment and exposure are estimated consistently, the TMLE estimator is asymptotically efficient. Additionally, its estimates will always stay within the bounds of the original outcome, thus making it more robust to outliers and spare data resulting in more reliable results. Finally, since TMLE fully incorporates machine learning, it is also a very attractive choice for analyzing complex observational data with a large number of variables and potentially complex relationships ^32^. While TMLE offers the most mature package for R users, researchers could also use one of the many causal inference packages available in python ^33^.

Assessing data for potential disparities is also important as datasets are increasingly utilized to build AI algorithms. Major companies including Google, Microsoft, and PwC have acknowledged that causal inference is an important framework for reviewing data and have updated their software packages to include tools for counterfactual modeling ^34–36^. Our open-source causal inference analytic pipeline is important not just for hospital quality metrics but also for researchers to identify biases in their datasets prior to developing models for prediction, classification and optimization.

Interestingly, our analysis found that all three of the treatments we assessed—IMV, RRT, and VP—did not lead to significant benefit in mortality outcomes across all race-ethnicities. There was a signal for potential harm in some patients treated with RRT, suggesting that critically ill patients may benefit while less ill patients may be harmed. Several recent study findings have shown that a “less is more” strategy may be beneficial in critical care ^37^ and further research is needed to understand which populations benefit most from life-sustaining interventions. This was beyond the scope of our current study, but sharing our repository publicly allows our analysis to be replicated and extended by researchers interested in exploring this question.

### Limitations

One of the limitations to our study is the categorization of patients into two broad racial-ethnic groups. We used this strategy due to the effective sample size once analysis is stratified across illness severity. In addition, the goal of this study was to demonstrate a framework for evaluating health disparities regardless of the axis of demography being investigated. Important differences in culture, ancestry, and lived experiences are lost when patients are aggregated into less precise groups, whether the comparison is for white vs. non-white, binary vs. non-binary, low- vs. high-income. Although we found no striking differences in treatments and outcomes between our two groups, disparities may be present in smaller subgroup analyses if the sample size had been larger. Even though MIMIC-IV contains 76,943 ICU stays and delineates 33 options for the race and ethnicity categories, power was not be enough to draw causal inferences on subpopulations. Furthermore, 17% (i.e., 3,983 out of 23,401) of patients in the dataset are categorized as “other,” highlighting the challenges of acquiring accurate demographic data at the bedside, particularly in a critically ill population. Further diversity, equity, and inclusion (DEI) efforts are needed at both the institutional and research level to improve our ability to collect and analyze disaggregated racial ethnic data ^38^. Other subgroup analyses, such as evaluating differences in treatments and outcomes across sex and gender identity, preferred language, and education and health literacy can also add to our understanding of how social determinants of health may lead to disparities.

## CONCLUSIONS

Causal inference enables us to explore the sources and drivers of disparities in healthcare, providing an opportunity for equality improvement initiatives or mitigate bias in AI algorithms. We provide an open-source analytic framework that allows healthcare providers, researchers, and policy makers to leverage a causal inference framework for health disparity and data science research.

## Data Availability

The data that support the study are available in MIMIC-IV with the identifier doi.org/10.1093/jamia/ocx084 publicly available on PhysioNet

https://physionet.org/content/mimiciv/2.2/

## ACKNOWLEDGEMENT

**Guarantor:** Struja

**Author Contributions:** Dr Struja and Mr. Matos designed the study and had full access to all the data in the study and take responsibility for the integrity of the data and the accuracy of the data analysis.

**Concept and design:** Struja, Celi, Matos, Jia, and Waschka.

**Acquisition, analysis, or interpretation of data:** All authors.

**Drafting of the manuscript:** Struja.

**Critical revision of the manuscript for important intellectual content:** All authors.

**Statistical analysis:** Struja, Cao, Waschka, and Matos.

**Administrative, technical, or material support:** Celi, Jia, and Waschka.

**Supervision:** Celi, Jia, and Waschka.

**Conflict of Interest Disclosures:** All authors confirm that they do not have a conflict of interest associated with this manuscript. YC and YJ are currently also affiliated with Verily life science, SSF, CA.

**Funding/Support:** This study was supported by unrestricted educational grants to Celi (NIBIB, R01 EB001659), Struja (Swiss National Science Foundation, P400PM_194497 / 1), Matos (Fulbright / FLAD Grant, Portugal, AY 2022/2023), and Sauer (German Research Foundation funded UMEA Clinician Scientist Program, FU356/12-2).

**Role of the Funder/Sponsor:** The funding sources had no role in the design and conduct of the study; collection, management, analysis, and interpretation of the data; preparation, review, or approval of the manuscript; and decision to submit the manuscript for publication.

**Availability of data:** The data that support the findings of this study are available in MIMIC-IV with the identifier doi.org/10.1093/jamia/ocx084. The database is publicly available on PhysioNet.

**Code availability:** The code that produces the results in this manuscript can be accessed at https://github.com/joamats/mit-tmle-sepsis, which includes detailed instructions for running the code.

## Supplementary Material

### Supplemental Online Content

**Table S1.** Comparison between target trial and the observational study of the effectiveness of invasive treatments in patients with sepsis (adapted from Hernán MA, NEJM 2021).

| Approach | Target Trial | Observational study |
| --- | --- | --- |
| Eligibility criteria | Patients with their first stay with least 24 hours stay fulfilling sepsis-3 criteria and $\geq 18$ years of age and known ethnicity | Same as for target trial |
| Treatment strategies | Initiation of mechanical ventilation, RRT, or vasopressors within the first 24 hours (72 hours for RRT) of ICU admissions | Same as for target trial |
| Treatment assignment | Intention-to-treat and per-protocol effects without blinding | Based on observed data |
| Outcomes | Primary: in-hospital death or discharge to hospice<br>Secondary: hospital-free days by day 28, nosocomial infections (CLABSI, CAUTI, SSI, VAP) | Same as for target trial |
| Follow-up | From treatment assignment until hospital day 30 | Same as for target trial |
| Estimation | Intention-to-treat and per-protocol effects | Intention-to-treat effect |

Abbreviations: RRT, renal replacement therapy; CLABSI, central-line associated bloodstream infection; CAUTI, catheter associated urinary tract infection; SSI, surgical site infection; VAP, ventilator associated pneumonia

**Table S2.**
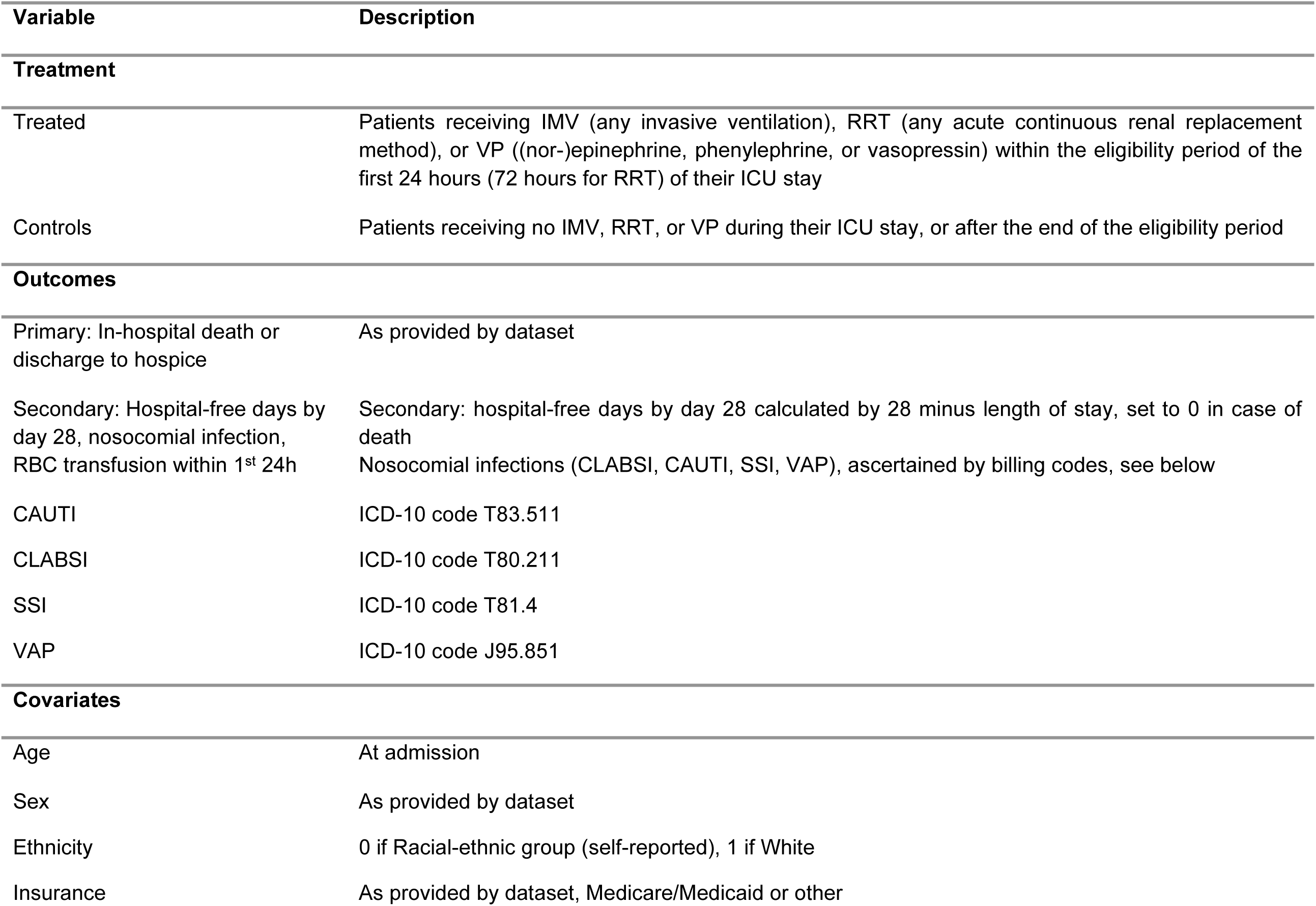
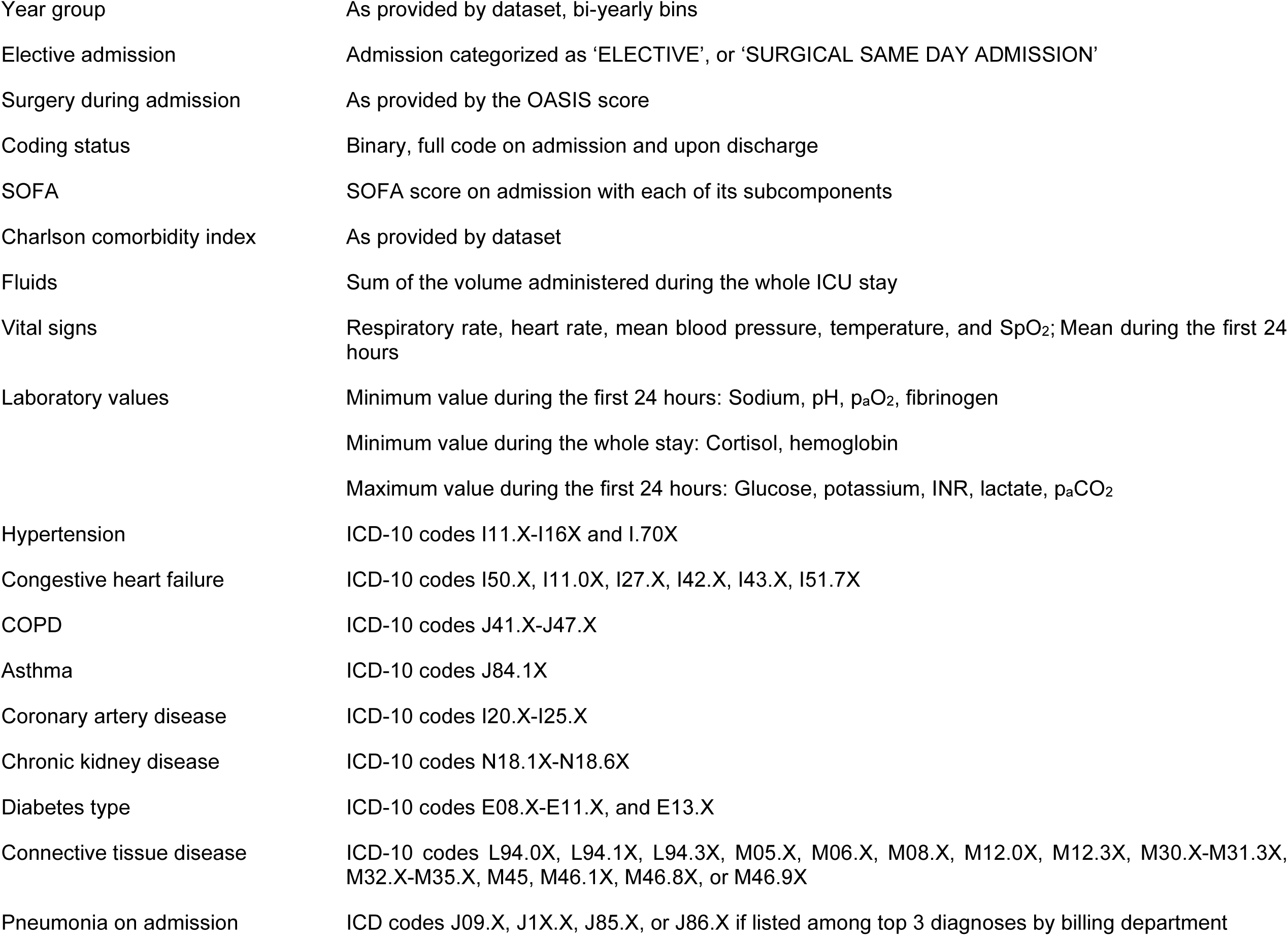
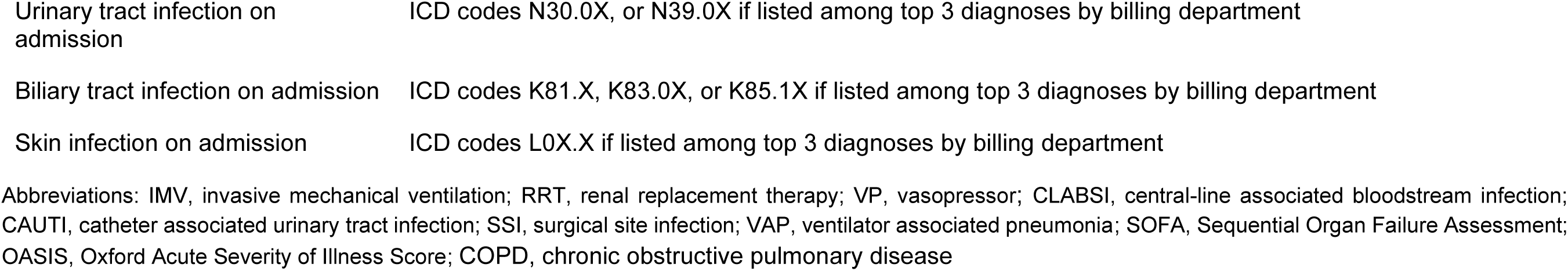
Variables used in the analysis to adjust the TMLE models.

**Table S3.**
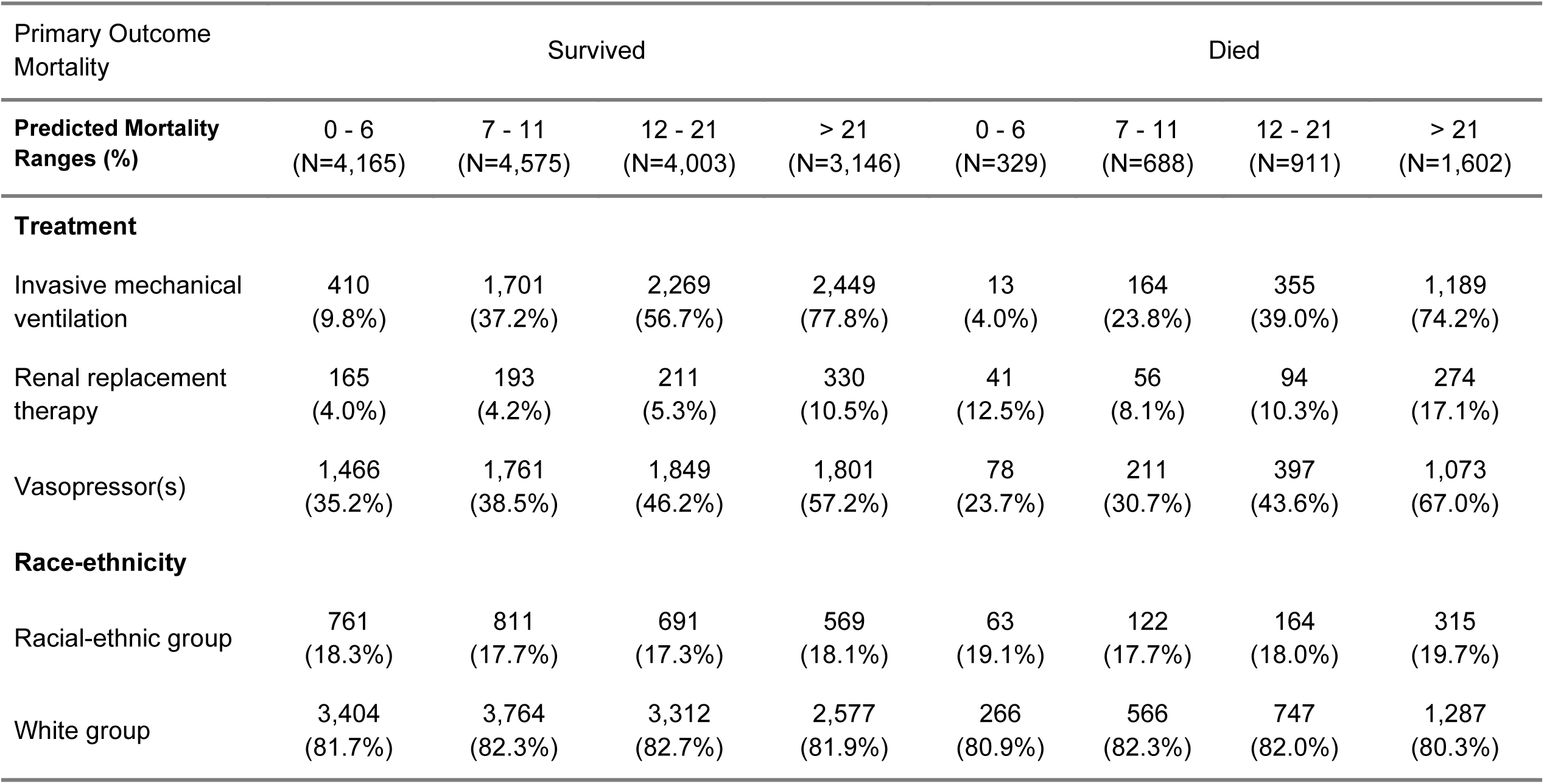
Positivity of mortality for every mortality probability, treatment, and race-ethnicity, to check for violations of the positivity assumption.

| Primary Outcome Mortality | Survived |  |  |  | Died |  |  |  |
| --- | --- | --- | --- | --- | --- | --- | --- | --- |
| Predicted Mortality Ranges (%) | 0 - 6<br>(N=4,165) | 7 - 11<br>(N=4,575) | 12 - 21<br>(N=4,003) | > 21<br>(N=3,146) | 0 - 6<br>(N=329) | 7 - 11<br>(N=688) | 12 - 21<br>(N=911) | > 21<br>(N=1,602) |
| **Treatment** |  |  |  |  |  |  |  |  |
| Invasive mechanical ventilation | 410<br>(9.8%) | 1,701<br>(37.2%) | 2,269<br>(56.7%) | 2,449<br>(77.8%) | 13<br>(4.0%) | 164<br>(23.8%) | 355<br>(39.0%) | 1,189<br>(74.2%) |
| Renal replacement therapy | 165<br>(4.0%) | 193<br>(4.2%) | 211<br>(5.3%) | 330<br>(10.5%) | 41<br>(12.5%) | 56<br>(8.1%) | 94<br>(10.3%) | 274<br>(17.1%) |
| Vasopressor(s) | 1,466<br>(35.2%) | 1,761<br>(38.5%) | 1,849<br>(46.2%) | 1,801<br>(57.2%) | 78<br>(23.7%) | 211<br>(30.7%) | 397<br>(43.6%) | 1,073<br>(67.0%) |
| **Race-ethnicity** |  |  |  |  |  |  |  |  |
| Racial-ethnic group | 761<br>(18.3%) | 811<br>(17.7%) | 691<br>(17.3%) | 569<br>(18.1%) | 63<br>(19.1%) | 122<br>(17.7%) | 164<br>(18.0%) | 315<br>(19.7%) |
| White group | 3,404<br>(81.7%) | 3,764<br>(82.3%) | 3,312<br>(82.7%) | 2,577<br>(81.9%) | 266<br>(80.9%) | 566<br>(82.3%) | 747<br>(82.0%) | 1,287<br>(80.3%) |

**Table S4.**
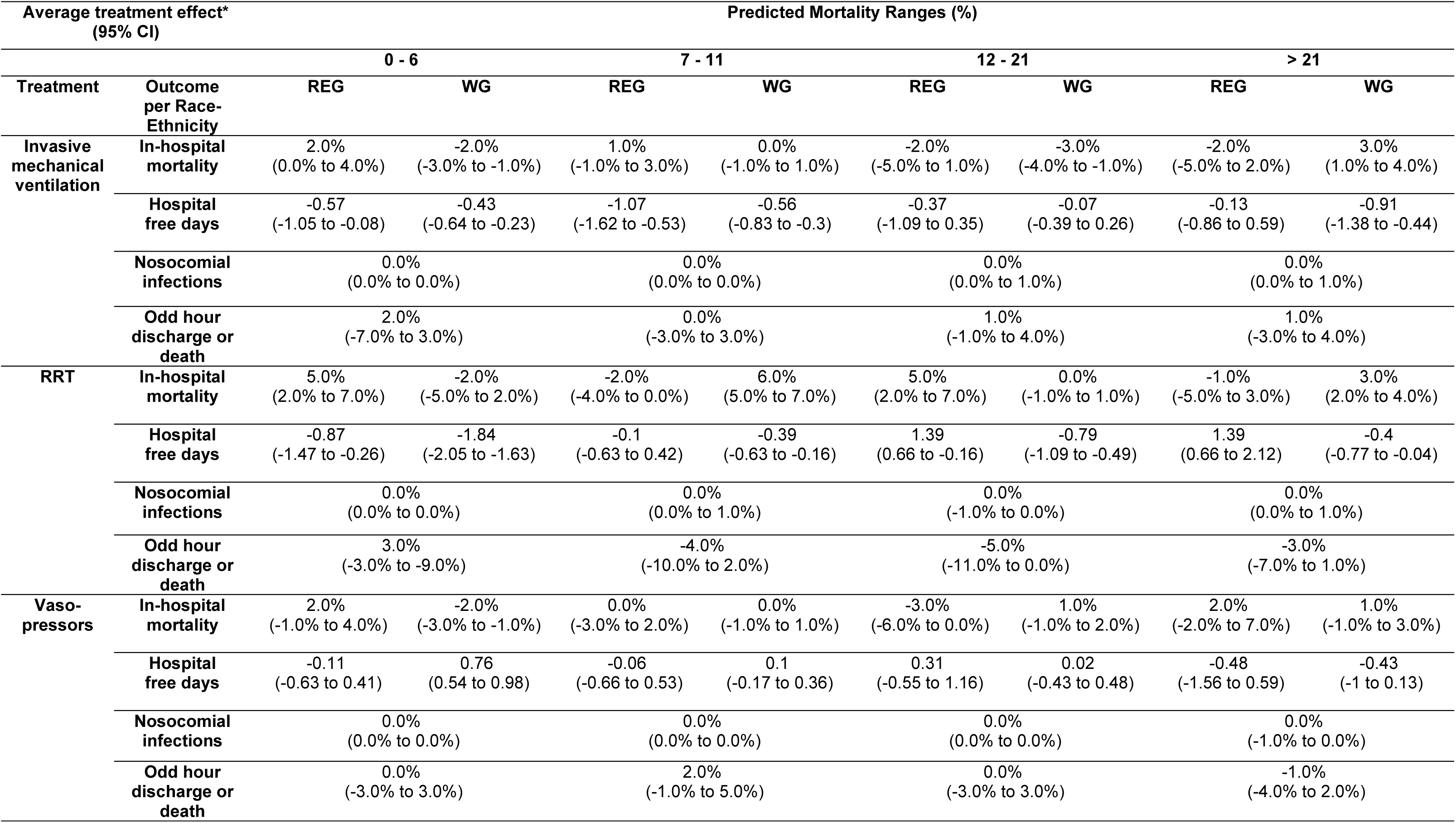
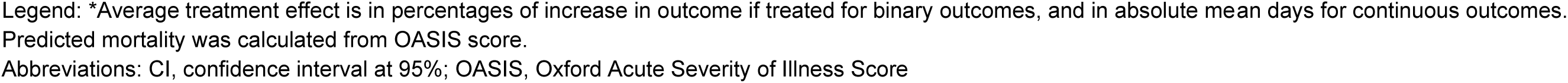
Primary and secondary outcomes using targeted maximum likelihood estimation.

**eTable 4:** Primary and secondary outcomes using targeted maximum likelihood estimation.
| Average treatment effect*<br>(95% CI) |  | Predicted Mortality Ranges (%) |  |  |  |  |  |  |  |
| --- | --- | --- | --- | --- | --- | --- | --- | --- | --- |
|  |  | 0 - 6 |  | 7 - 11 |  | 12 - 21 |  | > 21 |  |
| Treatment | Outcome<br>per Race-Ethnicity | REG | WG | REG | WG | REG | WG | REG | WG |
| Invasive<br>mechanical<br>ventilation | In-hospital<br>mortality | 2.0%<br>(0.0% to 4.0%) | -2.0%<br>(-3.0% to -1.0%) | 1.0%<br>(-1.0% to 3.0%) | 0.0%<br>(-1.0% to 1.0%) | -2.0%<br>(-5.0% to 1.0%) | -3.0%<br>(-4.0% to -1.0%) | -2.0%<br>(-5.0% to 2.0%) | 3.0%<br>(1.0% to 4.0%) |
|  | Hospital<br>free days | -0.57<br>(-1.05 to -0.08) | -0.43<br>(-0.64 to -0.23) | -1.07<br>(-1.62 to -0.53) | -0.56<br>(-0.83 to -0.3) | -0.37<br>(-1.09 to 0.35) | -0.07<br>(-0.39 to 0.26) | -0.13<br>(-0.86 to 0.59) | -0.91<br>(-1.38 to -0.44) |
|  | Nosocomial<br>infections | 0.0%<br>(0.0% to 0.0%) |  | 0.0%<br>(0.0% to 0.0%) |  | 0.0%<br>(0.0% to 1.0%) |  | 0.0%<br>(0.0% to 1.0%) |  |
|  | Odd hour<br>discharge or<br>death | 2.0%<br>(-7.0% to 3.0%) |  | 0.0%<br>(-3.0% to 3.0%) |  | 1.0%<br>(-1.0% to 4.0%) |  | 1.0%<br>(-3.0% to 4.0%) |  |
| RRT | In-hospital<br>mortality | 5.0%<br>(2.0% to 7.0%) | -2.0%<br>(-5.0% to 2.0%) | -2.0%<br>(-4.0% to 0.0%) | 6.0%<br>(5.0% to 7.0%) | 5.0%<br>(2.0% to 7.0%) | 0.0%<br>(-1.0% to 1.0%) | -1.0%<br>(-5.0% to 3.0%) | 3.0%<br>(2.0% to 4.0%) |
|  | Hospital<br>free days | -0.87<br>(-1.47 to -0.26) | -1.84<br>(-2.05 to -1.63) | -0.1<br>(-0.63 to 0.42) | -0.39<br>(-0.63 to -0.16) | 1.39<br>(0.66 to -0.16) | -0.79<br>(-1.09 to -0.49) | 1.39<br>(0.66 to 2.12) | -0.4<br>(-0.77 to -0.04) |
|  | Nosocomial<br>infections | 0.0%<br>(0.0% to 0.0%) |  | 0.0%<br>(0.0% to 1.0%) |  | 0.0%<br>(-1.0% to 0.0%) |  | 0.0%<br>(0.0% to 1.0%) |  |
|  | Odd hour<br>discharge or<br>death | 3.0%<br>(-3.0% to -9.0%) |  | -4.0%<br>(-10.0% to 2.0%) |  | -5.0%<br>(-11.0% to 0.0%) |  | -3.0%<br>(-7.0% to 1.0%) |  |
| Vaso-<br>pressors | In-hospital<br>mortality | 2.0%<br>(-1.0% to 4.0%) | -2.0%<br>(-3.0% to -1.0%) | 0.0%<br>(-3.0% to 2.0%) | 0.0%<br>(-1.0% to 1.0%) | -3.0%<br>(-6.0% to 0.0%) | 1.0%<br>(-1.0% to 2.0%) | 2.0%<br>(-2.0% to 7.0%) | 1.0%<br>(-1.0% to 3.0%) |
|  | Hospital<br>free days | -0.11<br>(-0.63 to 0.41) | 0.76<br>(0.54 to 0.98) | -0.06<br>(-0.66 to 0.53) | 0.1<br>(-0.17 to 0.36) | 0.31<br>(-0.55 to 1.16) | 0.02<br>(-0.43 to 0.48) | -0.48<br>(-1.56 to 0.59) | -0.43<br>(-1 to 0.13) |
|  | Nosocomial<br>infections | 0.0%<br>(0.0% to 0.0%) |  | 0.0%<br>(0.0% to 0.0%) |  | 0.0%<br>(0.0% to 0.0%) |  | 0.0%<br>(-1.0% to 0.0%) |  |
|  | Odd hour<br>discharge or<br>death | 0.0%<br>(-3.0% to 3.0%) |  | 2.0%<br>(-1.0% to 5.0%) |  | 0.0%<br>(-3.0% to 3.0%) |  | -1.0%<br>(-4.0% to 2.0%) |  |

Legend: \*Average treatment effect is in percentages of increase in outcome if treated for binary outcomes, and in absolute mean days for continuous outcomes. Predicted mortality was calculated from OASIS score. Abbreviations: CI, confidence interval at 95%; OASIS, Oxford Acute Severity of Illness Score

**Figure S1a.**
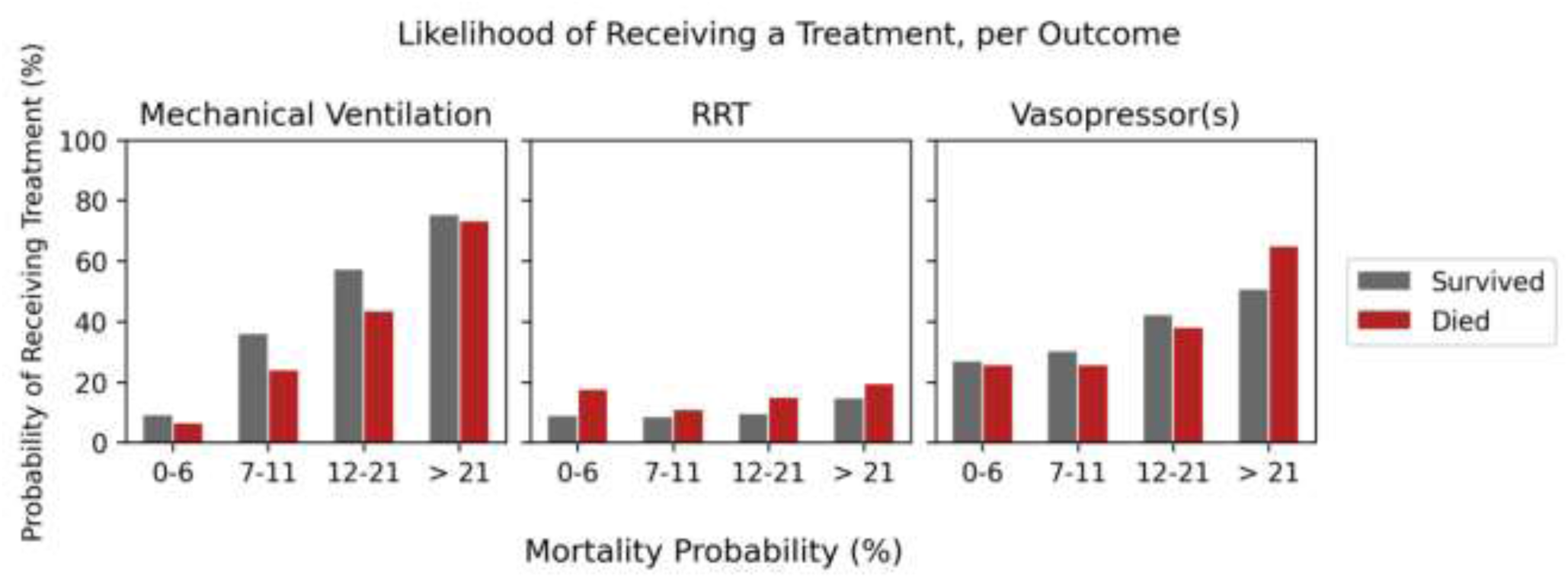
Distribution of patients across predicted mortality ranges, per invasive treatment for Racial-ethnic group only.

**Figure S1b.**
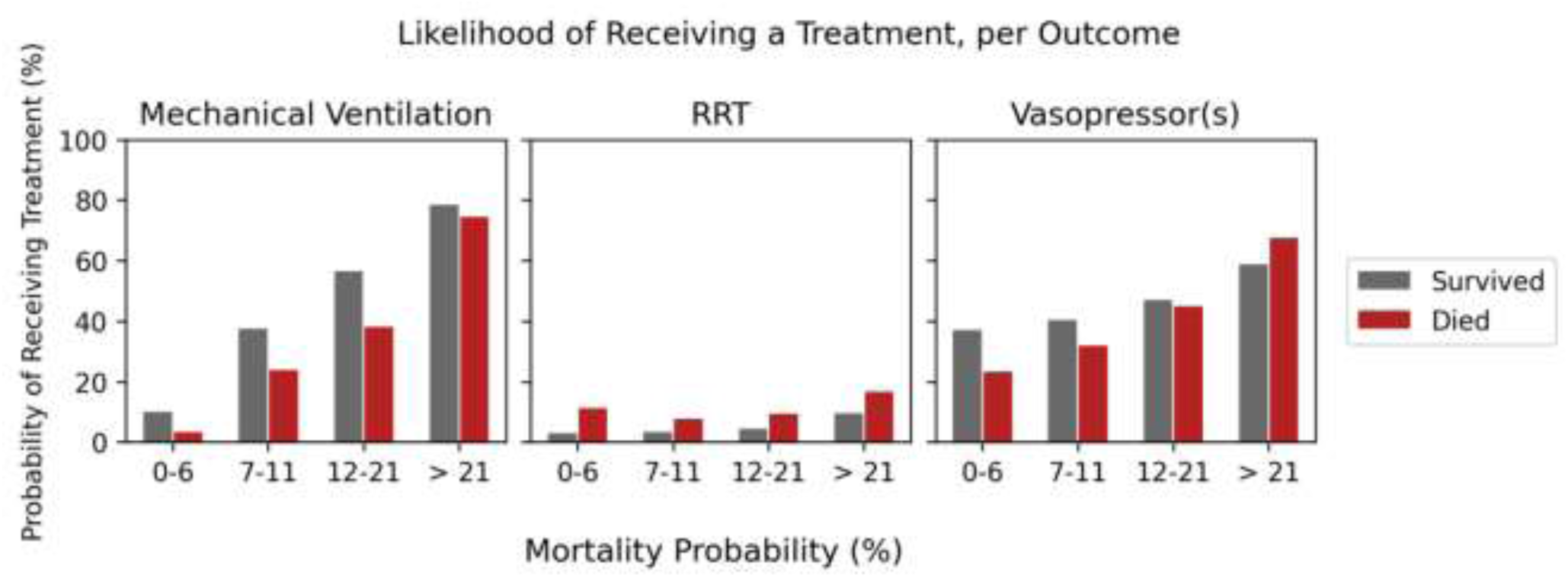
Distribution of patients across predicted mortality ranges, per invasive treatment for White group only. Abbreviations: RRT, renal replacement therapy

**Figure S2a.**
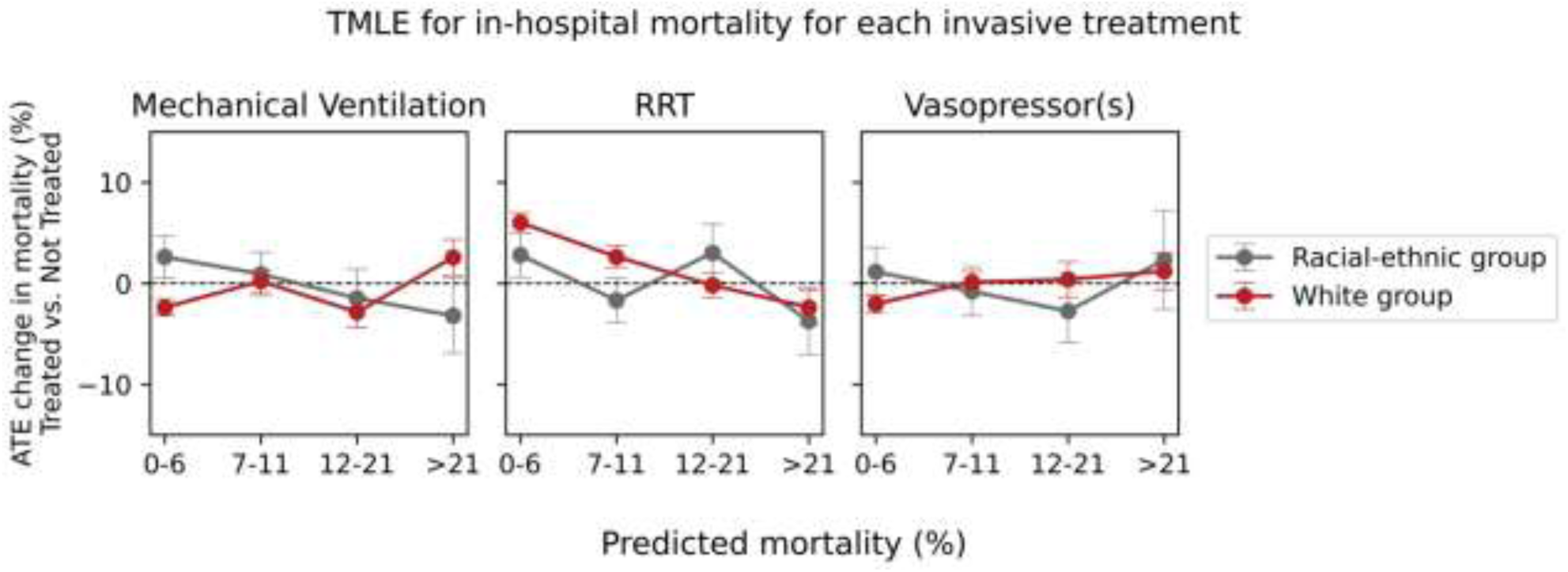
TMLE derived average treatment effects for change in probability of death over predicted mortality categories with an eligibility period for RRT of 1 day instead of 3 days. Abbreviations: TMLE, targeted maximum likelihood estimation; ATE, average treatment effects; RRT, renal replacement therapy

**Figure S2b.**
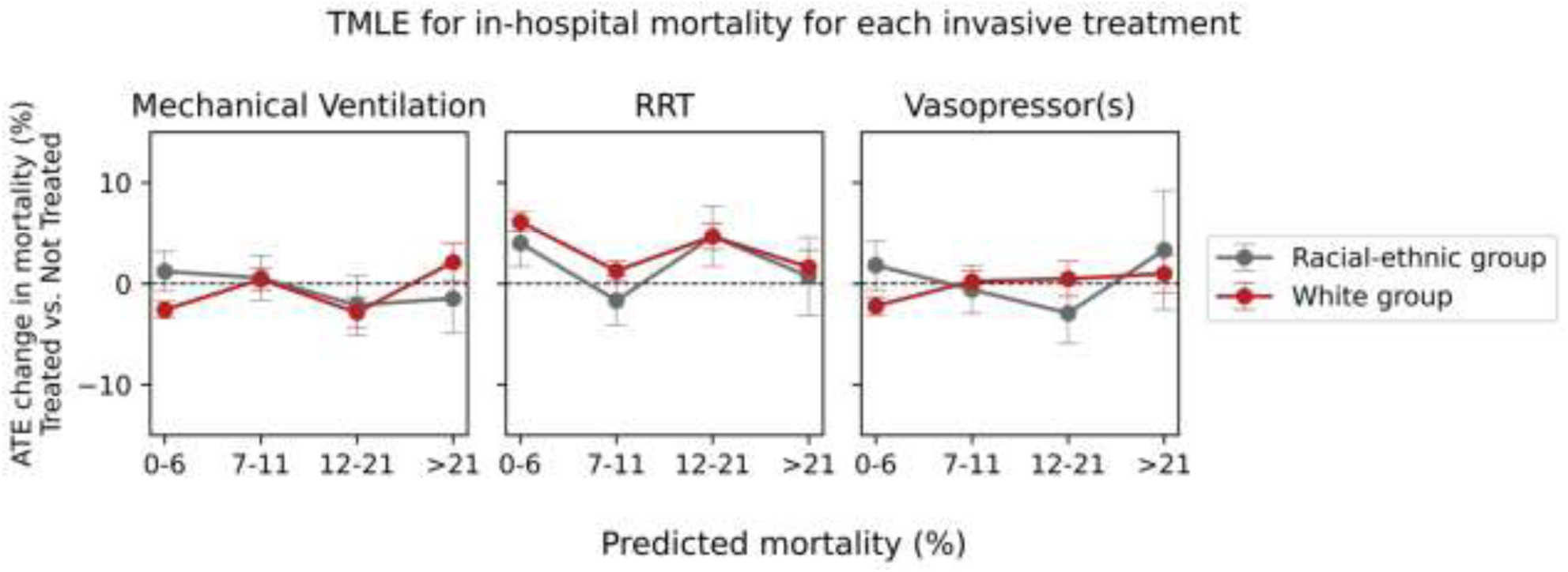
TMLE derived average treatment effects for change in probability of death over predicted mortality categories with an eligibility period for RRT of 5 days instead of 3 days.

**Figure S3.**
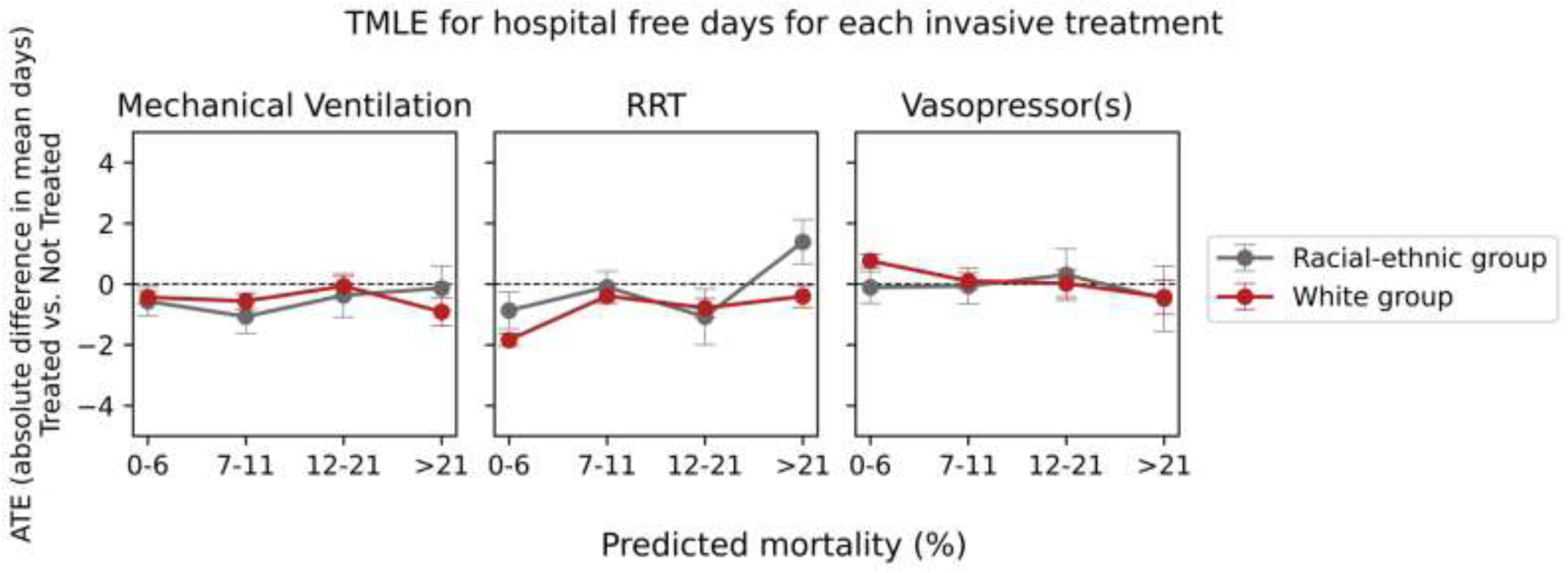
TMLE derived average treatment effects for change in mean hospital-free days over predicted mortality categories. Abbreviations: ATE, average treatment effects; TMLE, targeted maximum likelihood; RRT, renal replacement therapy

This supplemental material has been provided by the authors to give readers additional information about their work.

